# A comparative analysis of depression between pregnant and non-pregnant adolescents in a southwestern town in Nigeria

**DOI:** 10.1101/2022.04.01.22273317

**Authors:** Julianah T. Mosanya, Temilade A. Adegbite, Kazeem O. Adebayo, Bolaji E. Egbewale, Kayode T. Ijadunola

**Affiliations:** Department of Psychiatry, Osun State University (UNIOSUN) Teaching Hospital, Osogbo, Osun State, Nigeria; Department of Psychiatry, Lagos University Teaching Hospital, Lagos, Lagos State, Nigeria; Department of Psychiatry, Osun State University, Osogbo, Osun State, Nigeria; Department of Community Medicine, Ladoke Akintola University of Technology, Ogbomoso, Oyo State, Nigeria; Department of Community Health, Obafemi Awolowo University, Ile-Ife, Osun State, Nigeria

**Keywords:** Pregnant Adolescents, Depression, psychosocial, childhood physical abuse, childhood sexual abuse

## Abstract

**Background:** Adolescence constitutes a risk factor for mental health problems, and this may be further complicated by pregnancy. The rate of adolescent pregnancy is still extremely high in the sub-Saharan Africa including Nigeria. Pregnancy and mental health problems during adolescence constitute double vulnerability for negative outcomes for the adolescents and their offspring.

**Methodology:** The study was cross-sectional in design and it compared prevalence of depression and associated factors among pregnant and non-pregnant adolescents. It was conducted in Osogbo metropolis, Osun State, Southwest, Nigeria. The study population comprised pregnant adolescents (aged 15-19 years) attending antenatal care (ANC) in selected formal and informal health facilities. Non-pregnant adolescents who were equally attending services at the facilities were recruited as the control group. Information was obtained from the adolescents with the use of a structured questionnaire and data was analysed with IBM-SPSS version 21 software.

**Results:** Three hundred and thirty-four respondents (167 per group) were involved in the study; the pregnant adolescents had a mean age (±SD) of 17.92 (±1.13) years while the non-pregnant adolescents had a mean age of 17.70 (±1.23) years. The prevalence of depression among the pregnant adolescents was 8.4% while that of the non-pregnant adolescents was 3.0%. The result showed a statistically significant association between pregnancy status and depression among the adolescents (p= 0.033). Living arrangement was the only socio-demographic variable that had significant relationship with depression among the pregnant adolescents while living arrangement and employment status had significant relationships with depression among the non-pregnant adolescents. History of mental illness, childhood sexual abuse and anxiety symptoms showed significant relationship with depression among pregnant adolescents, however, only anxiety symptoms showed significant relationship with depression among non-pregnant adolescents.

**Conclusion:** The study concluded that the prevalence of depression is significantly higher among pregnant adolescents with similarities and differences in the factors associated with depression in the two groups.

## Introduction

The challenges of the developmental phase of adolescence are enormous and constitute risk for various mental health problems including depression. Depression is a common mental health disorder among adolescents (1), with equal incidence in males and females before puberty. However, there is a steady increase in the incidence in females after puberty till adulthood, when the risk doubles in them. (2). The vulnerability of females to depression beginning in adolescence (3) and the impact of an unidentified and untreated illness remain a challenge globally (4).

According to the World Health Organization, about 16 million girls aged 15-19 years and 1 million girls under 15 years give birth annually and most live in the low/middle-income countries (5).The burden of pregnancy or parenting during adolescence constitutes a risk for mental health problems in addition to the developmental risks of adolescence (6,7). Pregnancy and mental health problems in adolescents therefore constitute “double vulnerability” for negative outcomes in them and the future generation.

The poor mental health of the mothers has a potential to exert enormous impact on them, the unborn child and their infants after delivery. A pregnant woman with poor mental health is more likely to attend antenatal clinics poorly, comply poorly with treatment and subject herself to practices that may be detrimental to her and the foetus (7–9), consequently, impairing the success of public health interventions targeted at reduction of both maternal and neonatal morbidity and mortality. The two most important stages in human growth and development- the foetal period and adolescence are at stake in the context of adolescent pregnancy, especially when complicated by mental disorders. Poor attention to this vulnerable group of adolescents has a potential to set up a vicious cycle of poverty and mental health problems in them and the next generation (10).

Depression has been recognized as one of the most common perinatal mental health disorders among women including adolescent mothers. Despite its high prevalence, significance and availability of effective treatment options, it is usually undetected and untreated (6,11). Many studies on depression among adolescents have considered the school population extensively in both developed and developing countries (3,12–16). There are however fewer studies on depression among the pregnant adolescents and most of the available studies were conducted in the developed nations (17,18). The prevalence reported in some of these studies varied widely with rates as low as 1.6% (19) and as high as 68% (20), depending on various factors including the population, diagnostic criteria used, and age range of the respondents.

Commonly reported prevalence of current episode of depression among adolescents ranges between 2-10% (21–24). Although some studies in high-income countries showed that adolescent mothers are at a higher risk of depressive disorders compared with adult mothers (25,26) and non-pregnant adolescent girls (27), others refuted the evidence (28,29). However, there is a dearth of literature on both antepartum and postpartum mental disorders among adolescent girls in the developing countries where the burden of adolescent pregnancy is highest. This study aims to determine and compare the prevalence of depression and its relationship with sociodemographic and psychosocial variables between pregnant and non-pregnant adolescents in a south-western town in Nigeria.

## Materials And Methods

### Study setting and Participants

We employed a cross-sectional analytical design where comparison was made between the prevalence and correlates of depression among pregnant and non-pregnant adolescents aged 15 to 19 years. The participants were recruited from formal and informal health care facilities in Osogbo, the capital town of Osun State in south-west, Nigeria.

The pregnant adolescents were recruited from the public primary and secondary health care facilities as well as informal health facilities where they were attending antenatal care (ANC). Non-pregnant adolescents who attended the same facilities for one health reason or the other at the time of the study were recruited as the control group. Adolescents who were acutely ill or had chronic medical conditions were excluded from the study.

Simple random sampling method (balloting) was used to select two Primary Health Care Centres (PHC) from each of Osogbo and Olorunda Local Government areas in Osogbo town. The only secondary health facility in the town, the State Specialist Hospital, was also included (making a total of five formal health facilities). The secondary facility was included because health care services are free as obtainable in the PHCs. Two traditional / faith homes offering services that included antenatal care were also selected by snowballing method from each of the local governments as there was no sampling frame for this group. In total, nine facilities were included in the study.

Consecutive pregnant adolescents were recruited serially and the controls were also recruited from the same facilities; for formal health services, they were recruited from the general outpatient section while for the informal services, they were recruited among clients that came to receive care for other reasons besides pregnancy in the same facility. They were also recruited serially and matched for age of the pregnant adolescents, using frequency matching techniques.

### Sample size determination

Minimum sample size for each group required for this study was calculated using the formula for comparing two independent proportions (30).

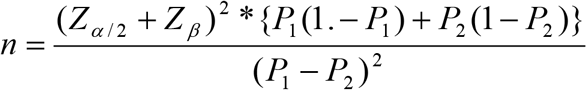

Where:

n = sample size in each group

Zα/2 = Standard normal deviate corresponding to confidence level; at confidence level of 95%, Z_α_ = 1.96 for a two tailed test

Z_β_ = Standard normal deviate corresponding to (1-Power); at power of 80%, Z_β_ = 0.84

P_1_ is the estimate of prevalence of depression in the pregnant adolescents

P_2_ is the estimate of prevalence of depression in the non-pregnant adolescents

P_1_ and P_2_ were derived from an earlier study in Brazil where 26.3% of pregnant adolescents had depression compared with 13.6% of the non-pregnant adolescents (27).

P_1_ = 0.263

P_2_ = 0.136

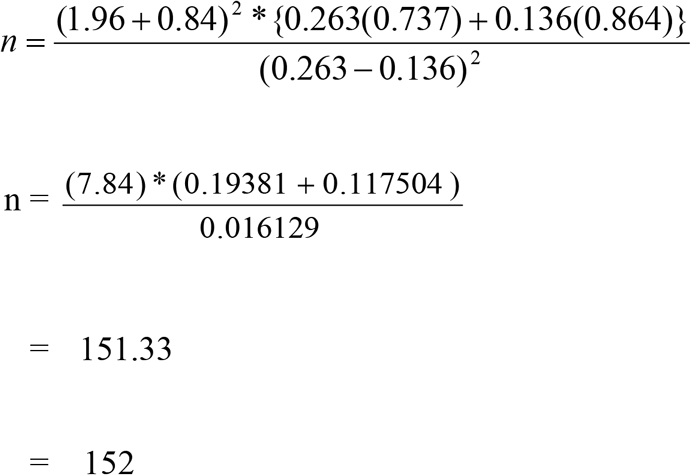

Ten percent was added to the sample size to make up for non-response (152 + 15) = 167 One hundred and sixty-seven respondents were recruited for each group in this study.

### Data Collection

Data was obtained from the respondents by means of a structured questionnaire. The questionnaire was administered to them after explaining the purpose of the research to them and obtaining informed consent/ assent. This was done after they had received care at the facility for the day. An identification sticker was placed on the hospital card of the patients that have been interviewed to avoid repeat sampling. The researcher ensured that there was privacy for the respondents while completing the questionnaire. The first section had questions on socio-demographic data, sexual history, whether pregnancy was planned and disposition to it. The other sections contained measures for depression, anxiety, social support, childhood sexual and physical abuse and recent abuse.

### Measures

#### Depression

The Beck Depression Inventory-II (BDI-II), which was used to assess depression in this study, is a popular screening instrument which has been validated and used previously among adolescents. It was adapted to the diagnostic criteria of major depressive episode of the Diagnostic and Statistical Manual of Mental Disorders-fourth edition (DSM-IV). It measures depressive symptomatology as well as assess the severity of depression in people who are thirteen years and above. It has 21 items, each item is scored 0 to 3 with possible total score of 0-63. A score of 0-16 is considered normal while 17-20, 21-30, and 31 and above are considered borderline, moderate and severe depression respectively Those who scored 21 and above were regarded as depressed because this cut-off has been reported previously to correlate significantly with clinical depression (31).

#### Anxiety

Patient Health Questionnaire for Depression and Anxiety-4 (PHQ-4): PHQ-4 combined both the two item Patient Health Questionnaire (PHQ-2) and the Generalized Anxiety Disorder (GAD-2). The scales were derived from the original eight and seven items PHQ-8 and GAD-7 respectively. PHQ-8 is used for screening for depression while GAD-7 is used for screening for anxiety. The first two of four items on PHQ-4 screened for anxiety and the last two screen for depression. Each item is rated on a scale of 0-3. Scores for each of the subscale is derived by adding scores for items on each subscales. Score ≥3 on each of the scale suggests that disorder. The scale for anxiety is used in this study as a measure of anxiety

#### Social Support

OSLO Social Support Questionnaire) has three items for assessing perceived social support, Minimum and maximum score obtainable on the scale are 3 and 14 respectively. Higher score denotes higher level of perceived social support. Scores of 3-8 is considered “poor social support”, 9-11 “moderate support and 12-14 “strong support”.

#### Childhood Physical and Sexual Abuse

Sexual and Physical Abuse Questionnaire (SPAQ) used in this study is a nine-item questionnaire that was designed as a screening tool for sexual and physical abuse in childhood. For this questionnaire, the operational definition of sexual and physical abuse involved having actual contact for sexual abuse, while presence of physical harm like bruises is required for physical abuse (32).

#### Abuse Assessment Screen (AAS)

Abuse assessment screen is a five-item screening tool for detection of domestic violence, an individual screens positive for abuse once any of the items is answered in the affirmative. The instrument was used as a measure of current/recent abuse in this study.

All instruments were translated to Yoruba and back translated to English to ensure content validity.

#### Statistical Analysis

Data were analyzed with the Statistical Package for Social Sciences (SPSS) version 21. Appropriate descriptive statistics such as percentages, means and standard deviation were used to summarize the data depending on the type of variable. Chi-square test was used to determine the relationship between the depression and the independent variables and significance level was set at p < 0.05.

#### Ethical Considerations

Ethical approval was obtained from the Institute of Public Health, Obafemi Awolowo University, Ile-Ife, Osun State. Permission was sought from the head of the selected health facilities and the doctors whose patients were involved in the research. Informed consent was obtained from the pregnant adolescents or parents/guardians. All pregnant adolescents gave informed consent irrespective of their age because they were considered “matured minors” (33). The parents/guardians of those that were less than 18 years old were required to give consent while the adolescents gave assent after which they were enrolled for participation. The questionnaire was administered to the adolescents privately. Privacy and confidentiality of all participants were ensured during and after data collection.

## Results

### Socio-demographic characteristics of pregnant and non-pregnant adolescents

Three hundred and thirty-four respondents (167 per group) were involved in the study. The mean (±SD) age of all the respondents was 17.81 (±1.18) years. There was no significant difference between the mean age of both groups of respondents (t = 1.670, p= 0.096). The pregnant adolescents had a mean age (±SD) of 17.92 (±1.13) years while the non-pregnant adolescents had a mean age of 17.70 (±1.23) years.

Thirty-nine percent of the pregnant adolescents compared with 84% of the non-pregnant adolescents were students and the difference was statistically significant (□^2^ = 77.24, p=0.001). Thirty seven percent of the non-pregnant adolescents compared with 11% of the pregnant ones had attained tertiary education and the difference was statistically significant (□^2^ = 29.525, p=0.001). Concerning employment status, 25% of the pregnant adolescents compared with 5% of the non-pregnant adolescents were employed. Although a higher proportion of pregnant adolescents were married (25%) compared with the non-pregnant ones (13%), the proportions of both groups that were never married were high at 72% and 84% respectively. Concerning living arrangements of the respondents, 77 (46%) pregnant adolescents compared with 127 (76%) non-pregnant ones lived with their parents. However, significantly higher proportions of the pregnant ones (29%) lived with a partner (husband or boyfriend) compared with the non-pregnant ones (6%), p<0.001. About 65% of the parents of the pregnant adolescents compared with 90.4% of the parents of the non-pregnant adolescents were married and the difference was statistically significant (□^2^ = 31.839, p=0.001). Likewise, 18% and 4.8% of mothers of the pregnant adolescents compared with 43.7% and 18.0% of the mothers of the non-pregnant ones had tertiary and postgraduate education respectively (□^2^ = 58.445, p=0.001). The pattern was also similar for the educational attainment of fathers of the respondents. (Table 1).

**Table 1:** Sociodemographic Characteristics of Pregnant and Non-pregnant Adolescents.

| **Variable** | **Pregnant**<br>**(n=167)**<br>**Frequency (%)** | **Non-pregnant**<br>**(n=167)**<br>**Frequency (%)** | **Total**<br>**(n=334)**<br>**Frequency(%)** | **Statistics** |
| --- | --- | --- | --- | --- |
| **Age (years)** |  |  |  |  |
| 15 | 7 (4.2) | 10 (6.0) | 17 (5.1) | $\chi^2 = 3.553$<br>df = 4<br>p = 0.470 |
| 16 | 17 (10.2) | 23 (13.8) | 40 (12.0) |  |
| 17 | 21 (12.6) | 28 (16.8) | 49 (14.7) |  |
| 18 | 60 (35.9) | 52 (31.1) | 112 (33.5) |  |
| 19 | 62 (37.1) | 54 (32.3) | 116 (34.7) |  |
| **Employment status** |  |  |  |  |
| Employed | 41(24.6) | 8 (4.8) | 49 (14.7) | $\chi^2 = 77.24$<br>df = 3<br>p = 0.001 |
| Unemployed | 29 (17.4) | 14 (8.4) | 43 (12.9) |  |
| Student | 65 (38.8) | 141 (84.4) | 206 (61.7) |  |
| Apprentice | 32 (19.2) | 4 (2.4) | 36 (10.7) |  |
| **Religion** |  |  |  |  |
| Christianity | 57 (34.1) | 80 (47.9) | 137 (41.0) | $\chi^2 = 7.061$ |
| Islam | 108 (64.7) | 84 (50.3) | 192 (57.5) | df =2 |
| Traditional | 2 (1.2) | 3 (1.8) | 5 (1.5) | **p = 0.029** |
| **Educational attainment** |  |  |  |  |
| Primary | 36 (21.6) | 22 (13.2) | 58 (17.4) | $\chi^2 = 29.525$ |
| Secondary | 107 (64.1) | 81 (48.5) | 188 (56.3) | df =3 |
| Tertiary | 19 (11.4) | 61 (36.5) | 80 (24.0) | **p = 0.001** |
| No formal | 5 (3.0) | 3 (1.8) | 8 (2.4) |  |
| **Marital status** |  |  |  |  |
| Never married | 120 (71.9) | 141 (84.4) | 261 (78.1) | $\chi^2 = 9.420$ |
| Married | 41 (24.6) | 22 (13.2) | 63 (18.9) | df =3 |
| Separated/divorced | 2 (1.2) | 0 (0) | 2 (0.6) | **p = 0.024** |
| Cohabiting | 4 (2.4) | 4 (2.4) | 8 (2.4) |  |
| **Living arrangement** |  |  |  |  |
| Partner | 49 (29.3) | 10 (6.0) | 59 (17.7) | $\chi^2 = 55.329$ |
| Parents | 77 (46.1) | 127 (76.0) | 204 (61.1) | df =3 |
| Grandparents | 34 (20.4) | 11 (6.6) | 45 (13.5) | **p = 0.001** |
| Alone | 7 (4.2) | 19 (11.4) | 26 (7.8) |  |

### Psychosocial characteristics of pregnant and non-pregnant adolescents

The proportion of those that signified that they have had history of mental illness was 5.4% among the pregnant adolescents compared with 1.8% of the non-pregnant peers, the difference was not statistically significant (p=0.139).

Twenty-three (13.8%) of the pregnant adolescents compared with 30 (18.0%) of the non-pregnant adolescents had poor social support (p=0.084). Twenty-five (15%) and 33 (19.8%) of the pregnant adolescents compared with 16 (9.6%) and 30 (18.0%) of non-pregnant adolescents had history of childhood physical and sexual abuse respectively, but the difference in proportions were not statistically significant (p=0.133 and 0.675). Similarly, the proportions of those exposed to recent abuse was comparable between the two groups (p=0.082). Fewer, 13 (7.8%) of the pregnant adolescents compared with 24 (14.4%) of the non-pregnant adolescents screened positive for anxiety, but the difference did not reach statistical significance (p= 0.055) (Table 2).

**Table 2:**
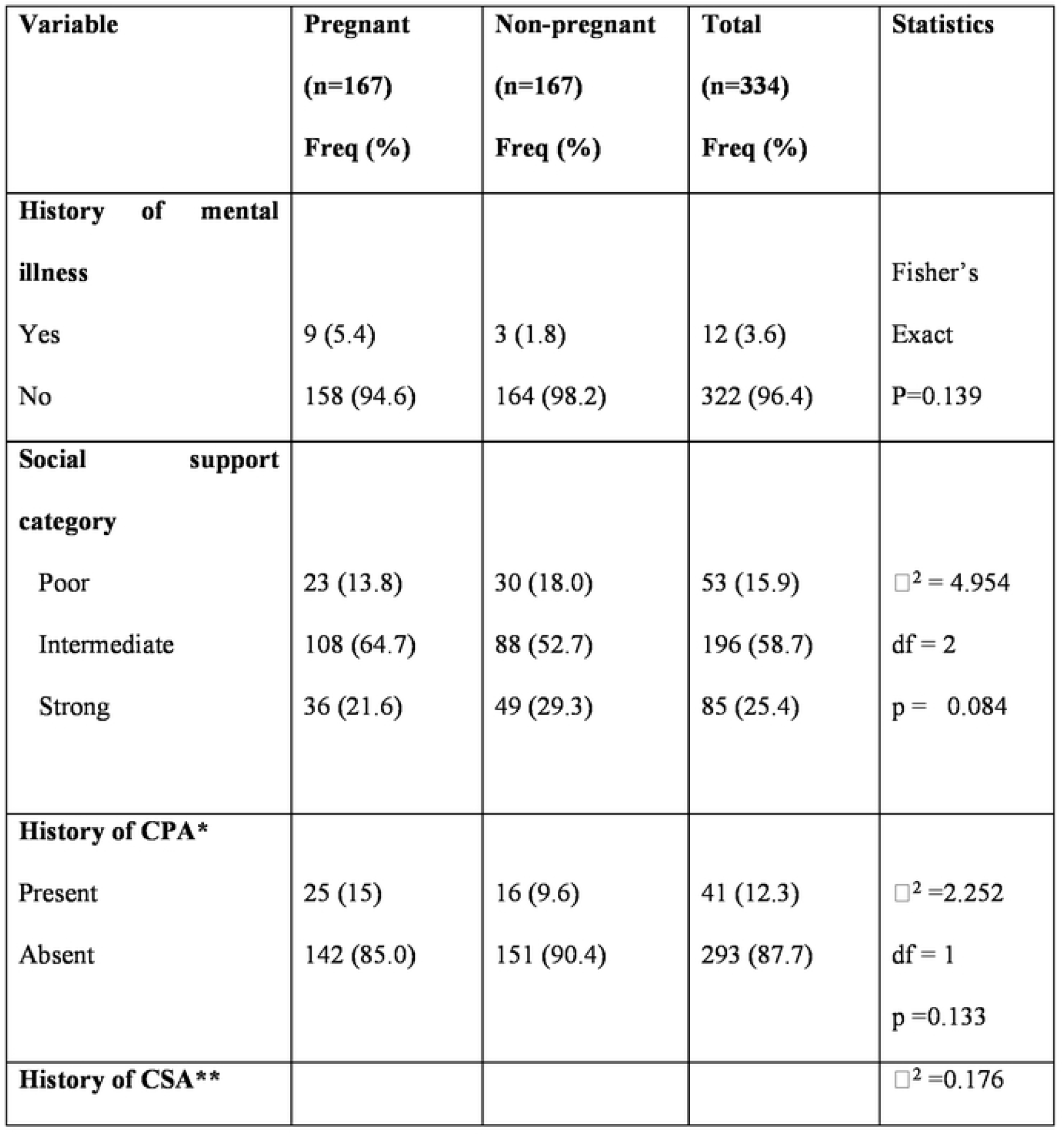
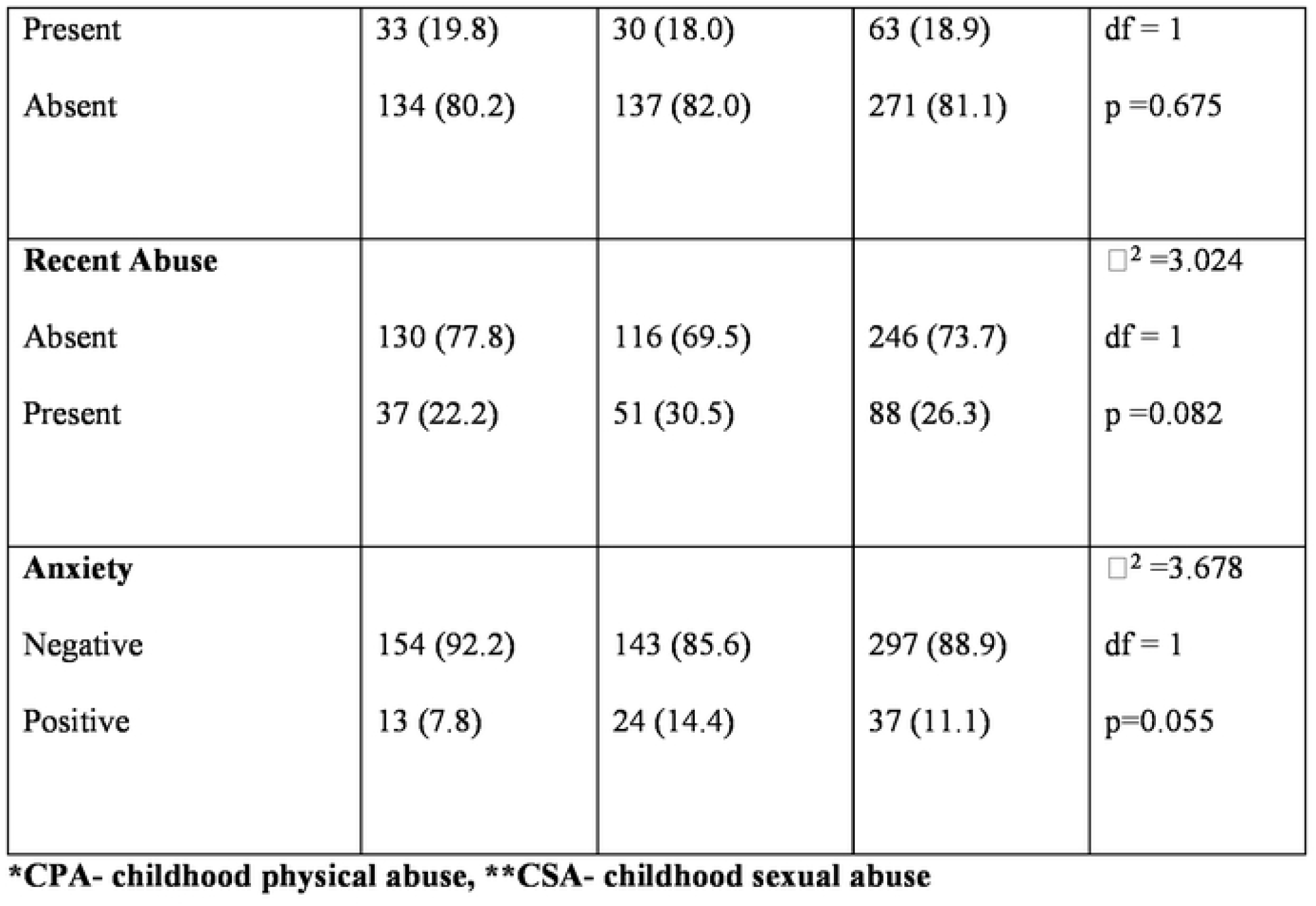
Psychosocial Variables of Pregnant and Non-pregnant Adolescents.

| **Variable** | **Pregnant**<br>**(n=167)**<br>**Freq (%)** | **Non-pregnant**<br>**(n=167)**<br>**Freq (%)** | **Total**<br>**(n=334)**<br>**Freq (%)** | **Statistics** |
| --- | --- | --- | --- | --- |
| **History of mental illness** |  |  |  | Fisher's |
| Yes | 9 (5.4) | 3 (1.8) | 12 (3.6) | Exact |
| No | 158 (94.6) | 164 (98.2) | 322 (96.4) | P=0.139 |
| **Social support category** |  |  |  |  |
| Poor | 23 (13.8) | 30 (18.0) | 53 (15.9) | $\chi^2 = 4.954$ |
| Intermediate | 108 (64.7) | 88 (52.7) | 196 (58.7) | df = 2 |
| Strong | 36 (21.6) | 49 (29.3) | 85 (25.4) | p = 0.084 |
| **History of CPA\*** |  |  |  |  |
| Present | 25 (15) | 16 (9.6) | 41 (12.3) | $\chi^2 = 2.252$ |
| Absent | 142 (85.0) | 151 (90.4) | 293 (87.7) | df = 1 |
|  |  |  |  | p = 0.133 |
| **History of CSA\*\*** | | | | $\chi^2 = 0.176$ |
| Present | 33 (19.8) | 30 (18.0) | 63 (18.9) | df = 1 |
| Absent | 134 (80.2) | 137 (82.0) | 271 (81.1) | p = 0.675 |
| **Recent Abuse** | | | | $\chi^2 = 3.024$ |
| Absent | 130 (77.8) | 116 (69.5) | 246 (73.7) | df = 1 |
| Present | 37 (22.2) | 51 (30.5) | 88 (26.3) | p = 0.082 |
| **Anxiety** | | | | $\chi^2 = 3.678$ |
| Negative | 154 (92.2) | 143 (85.6) | 297 (88.9) | df = 1 |
| Positive | 13 (7.8) | 24 (14.4) | 37 (11.1) | p = 0.055 |

\*CPA- childhood physical abuse, \*\*CSA- childhood sexual abuse

### Planning status of pregnancy and its disposition among pregnant adolescents

Figure 1 and 2 shows the planning status of the pregnancy and their disposition to it among pregnant adolescents.

**Figure 1:**
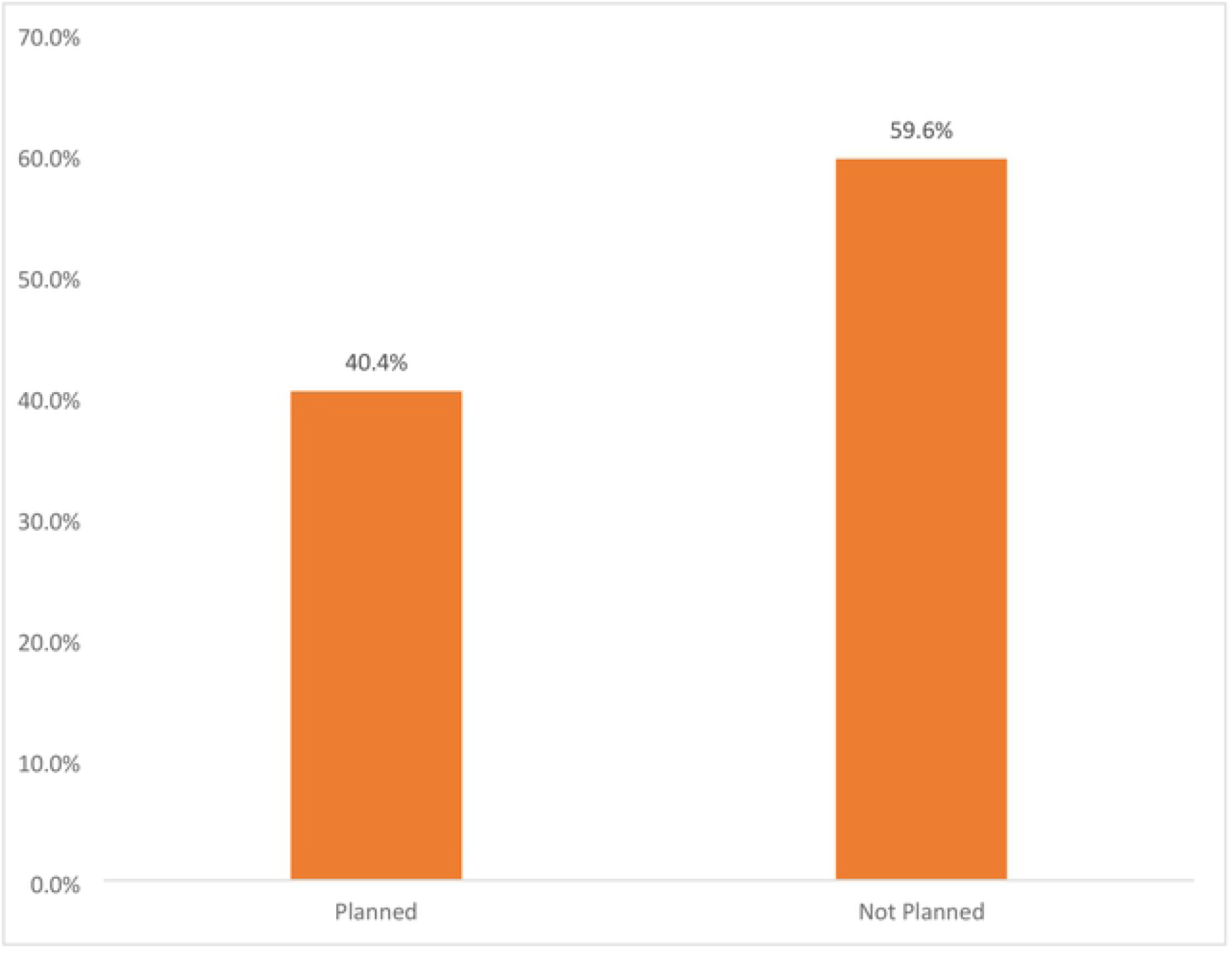
Planning Status of the Pregnancies of the Pregnant Adolescents.

**Figure 2:**
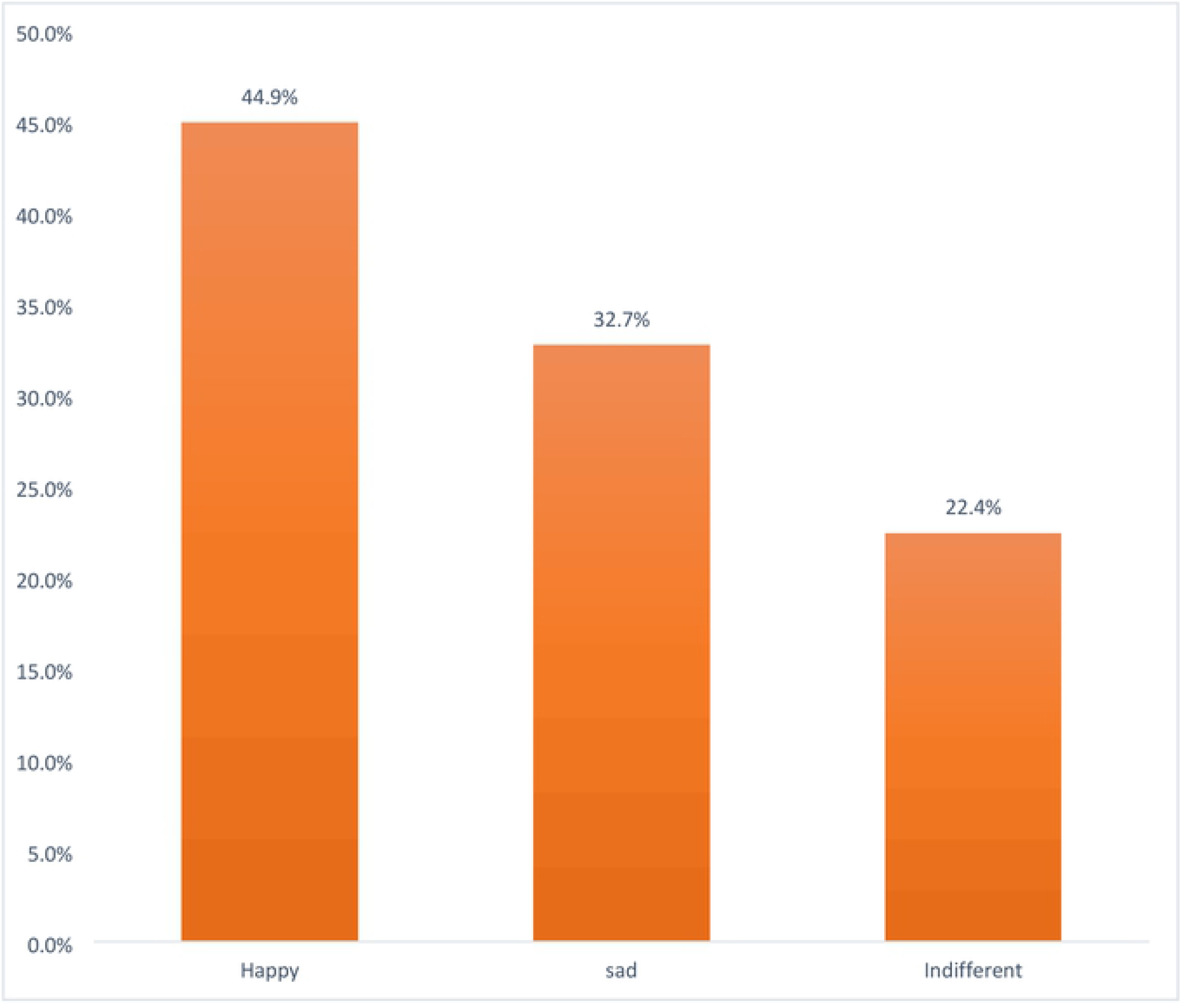
Disposition of Pregnant Adolescents in Osogbo towards their Pregnancies.

### Depression

The mean BDI score (which assessed depression among the respondents) of the pregnant adolescents was significantly higher, 8.32 (±8.17) than that of the non-pregnant adolescents (6.71 (±7.85) (p<0.001). Overall, 19 (5.7%) of the respondents were categorized as being depressed, 14 (8.4%) among the pregnant adolescents and 5 (3.0%) among the non-pregnant adolescents. The proportion of pregnant adolescents categorized as depressed was significantly higher than that of the non-pregnant adolescents (p= 0.033).

### Relationship between depression and sociodemographic and Psychosocial variables among the respondents

Table 3 shows the relationship between depression and selected sociodemographic variables among pregnant and non-pregnant adolescents. Living arrangement was the only variable that had significant relationship with depression among the pregnant adolescents while living arrangement and employment status had significant relationships with depression among the non-pregnant adolescents.

**Table 3:** Relationship between depression and socio-demographic variables in pregnant and non-pregnant adolescents.

| Pregnant adolescents | Non-pregnant adolescents |
| --- | --- |

| Variable | Not<br>Depressed<br>(n=153)<br>Freq (%) | Depressed<br>(n=14)<br>Freq (%) | Total<br>(n=167)<br>Freq (%) | Statistics | Not<br>Depressed<br>(n=162)<br>Freq (%) | Depressed<br>(n=5)<br>Freq (%) | Total<br>(n=167)<br>Freq (%) |
| --- | --- | --- | --- | --- | --- | --- | --- |
| **Marital**<br>**Status** |  |  |  | Fisher's |  |  |  |
| Married | 38 (92.7) | 3 (7.3) | 41 (100.0) | Exact | 21 (95.5) | 1 (4.5) | 22 (100.0) |
| Not married | 115 (91.3) | 11 (8.7) | 126 (100.0) | P=1.000 | 141 (97.2) | 4 (2.8) | 145 (100.0) |
| **Educational**<br>**attainment** |  |  |  | Fisher's |  |  |  |
| <Secondary | 37(90.2) | 4 (9.8) | 41 (100.0) | Exact | 25(100.0) | 0 (0.0) | 25 (100.0) |
| ≥Secondary | 116 (92.1) | 10 (7.9) | 126(100.0) | P=0.748 | 137 (96.5) | 5 (3.5) | 142(100.0) |
| **Employment**<br>**Status** |  |  |  |  |  |  |  |
| Employed | 38 (92.7) | 3 (7.3) | 41 (100.0) | $\chi^2=0.814^*$ | 6 (75.0) | 2 (25.0) | 8 (100.0) |
| Unemployed | 27 (93.1) | 2 (6.9) | 29 (100.0) | df=3 | 13 (92.9) | 1 (7.1) | 14 (100.0) |
| Student | 60 (92.3) | 5 (7.7) | 65 (100.0) | p=0.846 | 140(99.3) | 1 (0.7) | 141 (100.0) |
| Apprentice | 28 (87.5) | 4 (12.5) | 32 (100.0) |  | 3 (75.0) | 1 (25.0) | 4 (100.0) |
| **Religion** |  |  |  |  |  |  |  |
| Christianity | 53 (93.0) | 4 (7.0) | 57 (100.0) | $\chi^2=2.507^*$ | 77 (96.3) | 3 (3.8) | 80 (100.0) |
| Islam | 99 (91.7) | 9 (8.3) | 108 (100.0) | df =2 | 82 (97.6) | 2 (2.4) | 84 (100.0) |
| Traditional | 1 (50.0) | 1 (50.0) | 2 (100.0) | p=0.286 | 3(100.0) | 0 (0.0) | 3 (100.0) |
| **Living arrangement** |  |  |  |  |  |  |  |
| Partner | 46 (93.9) | 3 (6.1) | 49 (100.0) | $\chi^2$ | 9 (90.9) | 1(10.0) | 10(100.0) |
| Parents | 71 (92.2) | 6 (7.8) | 77 (100.0) | =12.903* | 126 (99.2) | 1 (0.8) | 127 (100.0) |
| Grandparents | 33 (97.1) | 1 (2.9) | 34 (100.0) | df =3 | 11 (100.0) | 0(0.0) | 11 (100.0) |
| Alone | 3 (42.9) | 4 (57.1) | 7 (100.0) | p=**0.005** | 16 (84.2) | 3 (15.8) | 19 (100.0) |
| **Parent's Marital Status** |  |  |  | Fisher's |  |  |  |
| Married | 98 (90.7) | 10 (9.3) | 108 (100.0) | Exact | 146 (96.7) | 5 (3.3) | 151 (100.0) |
| Not Married | 55 (93.2) | 4 (6.8) | 59 (100.0) | P=0.772 | 16 (100.0) | 0 (0.0) | 16 (100.0) |

Table 4 shows the relationship between depression and psychosocial variables among pregnant and non-pregnant adolescents. Among pregnant adolescents, 33% of those with positive history of mental illness compared with only 7% of those with negative history of mental illness had depression and the difference was statistically significant (Fisher’s Exact Test, p=0.029). Similarly, a higher proportion of those who were exposed to childhood sexual abuse (18%) compared to those who were not exposed (6%) had depression (p=0.023). Conversely, exposure to childhood physical abuse had no significant relationship with depression among the pregnant adolescent. Also, social support and current abuse had no relationship with depression status. Similarly, whether pregnancy was planned or not as well as disposition to the pregnancy showed no relationship with depression among them. Anxiety however showed significant relationship with depression as significantly higher proportions of those that screened positive for anxiety (38.5%) compared with those that were negative (5.8%) had depression (p=0.001).

**Table 4:** Relationship between depression and psychosocial variables in pregnant and non-pregnant adolescents.

| Pregnant adolescents | Non-pregnant adolescents |
| --- | --- |

| Variable | Not<br>Depressed<br>(n=153)<br>Freq (%) | Depressed<br>(n=14)<br>Freq (%) | Total<br>(n=167)<br>Freq (%) | Statistics | Not<br>Depressed<br>(n=162)<br>Freq (%) | Depressed<br>(n=5)<br>Freq (%) | Total<br>(n=167)<br>Freq (%) |
| --- | --- | --- | --- | --- | --- | --- | --- |
| **Hx of mental illness** |  |  |  | Fisher's<br>Exact<br>**P=0.029** | 3(100.0) | 0 (0.0) | 3(100.0) |
| Yes | 6 (66.6) | 3 (33.3) | 9 (100.0) |  | 159 (97.0) | 5(3.0) | 164 (100.0) |
| No | 147 (93.0) | 11 (7.0) | 158 (100.0) |  |  |  |  |
| **Social support** |  |  |  |  |  |  |  |
| Poor | 20 (87.0) | 3 (13.0) | 23 (100.0) | $\chi^2=4.954$ | 28 (93.3) | 2(6.7) | 30 (100.0) |
| Intermediate | 99(91.7) | 9 (8.3) | 108 (100.0) | df=2 | 87 (98.8) | 1 (1.1) | 88 (100.0) |
| Strong | 34 (94.4) | 2 (5.6) | 36(100.0) | p=0.611 | 47 (95.9) | 2(4.1) | 49 (100.0) |
| **Hx of CPA\*** |  |  |  |  |  |  |  |
| Absent | 131(92.3) | 11 (7.7) | 142 (100.0) | Fisher's | 147(97.4) | 4 (2.6) | 151 (100.0) |
| Present | 22 (88.0) | 3 (12.0) | 25 (100.0) | Exact<br>p=0.443 | 15 (93.8) | 1 (6.3) | 16 (100.0) |
| **Hx of CSA++** |  |  |  |  |  |  |  |
| Absent | 126 (94.0) | 8 (6.0) | 134 (100.0) | $\chi^2=5.141$ | 134 (97.8) | 3 (2.2) | 137 (100.0) |
| Present | 27 (81.8) | 6 (18.2) | 33 (100.0) | df=1 | 28 (93.3) | 2(6.7) | 30 (100.0) |
|  |  |  |  | **p=0.023** |  |  |  |
| **Recent Abuse** |  |  |  |  |  |  |  |
| Absent | 118 (90.8) | 12 (9.2) | 130 (100.0) | Fisher's | 113 (97.4) | 3 (2.6) | 116 (100.0) |
| Present | 35 (94.6) | 2 (5.4) | 37 (100.0) | Exact | 49 (96.1) | 2 (3.9) | 51 (100.0) |
|  |  |  |  | **p =0.737** |  |  |  |
| **Anxiety** |  |  |  |  |  |  |  |
| Negative | 145 (94.2) | 9 (5.8) | 144 (100.0) | $\chi^2 = 16.606$ | 141 (98.6) | 2 (1.4) | 143 (100.0) |
| Positive | 8 (61.5) | 5 (38.5) | 13 (100.0) | df = 1 | 21 (87.5) | 3 (12.5) | 24 (100.0) |
|  |  |  |  | **p=0.001** |  |  |  |
| **Pregnancy\*+ status** | | | | $\chi^2 = 0.137$ | | | |
| Planned | 62 (92.5) | 5 (7.5) | 67 (100.0) | df = 1 |  |  |  |
| Not planned | 90 (90.9) | 9 (9.1) | 99 (100.0) | p=0.711 | ----- | ----- | ----- |
| **Pregnancy disposition<sup>s</sup>** |  |  |  |  |  |  |  |
| Happy | 69 (93.2) | 5 (6.8) | 74 (100/0) | $\chi^2$ | | | |
| Sad | 47 (87.0) | 7 (13.0) | 54 (100.0) | =2.035** | ----- | ----- | ----- |
| Indifferent | 35 (94.6) | 2 (5.4) | 37 (100.0) | df = 2 |  |  |  |
|  |  |  |  | **p=0.362** |  |  |  |

\*CPA- childhood physical abuse; ++CSA-childhood sexual abuse; \*\*likelihood ratio $\chi^2$
\*+n=166 ; <sup>s</sup>n=165

Among non-pregnant adolescents, the history of mental illness, social support, childhood physical as well as sexual abuse had no relationship with depression. Current experience of abuse also had no relationship with depression. However, screening positive for anxiety showed significant relationship with depression with 12.5% of those with anxiety compared with 1.4% of those without anxiety having depression, p=0.022.

## DISCUSSION

The prevalence of depression among the pregnant adolescents (8.4%) was significantly higher than that of the non-pregnant adolescents (3.0%), p=0.033, this pattern followed the previous findings of pregnant adolescents having a higher risk of depression than their non-pregnant counterparts (27). The higher prevalence in the pregnant adolescents as found in this study may be due to the stress and/or physiological changes of pregnancy and social discrimination associated with teenage pregnancy (34). It may also be that the presence of depression actually predisposed to pregnancy in the adolescents as earlier studies suggested, that depression predisposes the adolescents to high-risk sexual behaviour which may increase the incidence of pregnancy in them and may account for the increased prevalence of depression among the pregnant adolescents (35,36). This finding highlights the importance of paying close attention to the mental health of adolescents in general and the pregnant ones in particular.

This observed relationship between studentship status and depression could make one to consider the possibility that being a student is protective against depression or that those who were not students had other risk factors like poverty that increased the risk of depression in them. On the one hand, it is possible that the presence of depressive disorder in these other groups predisposed them to dropping out of school, thereby looking for other things to do beyond schooling. On the other hand, depression in them could be a psychological reaction to dropping out of school which was informed by reasons out of their control. There may be need to watch out for depression in adolescents who are out of school. This finding is in agreement with a study in India that found a significantly higher prevalence of depression among female adolescents that dropped out of school (19).

Concerning the relationship of depression with living arrangements in this study, the proportion of respondents who stayed alone or with partners (boyfriend or husbands) who had depression was significantly higher than those who stayed with their parents or grandparents (p<0.001). This finding suggests that living with parents or grandparents may be a protective factor against depression in this age group rather than living alone or living with a partner. Furthermore, living with partner, especially when unmarried may put significant emotional strain on those who were practicing it, therefore increasing the risk of depression in them since the practice is not culturally acceptable in the environment where the study was carried out. Beyond this, the social support available for those living alone or with partners may not be as good as those living with their parents or grandparents among this population, thereby putting significant strain on their emotional health. The finding of an association of depression with few socio-demographic factors in this study is contrary to the finding of an earlier study that found no such association in pregnant adolescents (37).

Among the psychosocial factors examined, three factors; the past history of mental illness, anxiety and childhood sexual abuse had a significant relationship with depression. The past history of mental illness having a significant relationship with depression among the pregnant adolescents is not surprising because depression and different other mental illnesses are chronic illnesses that are prone to relapse, so anyone who has had an episode could have further episodes (38). The fact that there is no significant relationship between past history of mental illness and depression among the non-pregnant adolescents could then suggest that there is a relationship between mental illness and poor reproductive behaviour as suggested in previous studies (39), which could have predisposed the pregnant adolescents to the pregnancy in the first place. The type of mental illness that those with history of mental illness had previously was however not documented in this study, so it is not clear whether the past mental illness they had was depression or not.

The finding of a relationship between depression and childhood sexual abuse but not with childhood physical abuse among the pregnant adolescents in this study is surprising, since they are both forms of violence. It therefore raises the question as to whether sexual abuse has more emotional impact on the victims than physical abuse. In addition, one may want to know the contribution of sexual abuse to the occurrence of pregnancy as previous researches suggested (40). The finding of an association between depression and childhood sexual abuse in this study is in agreement with extant literature (41,42). Presence of anxiety symptoms had significant relationship with depression, this further confirms the strong possibility of depression co-existing with other psychological illnesses, especially anxiety symptoms/disorders as previous studies have documented. It is interesting to note that neither planned status of pregnancy nor disposition to the pregnancy had significant relationship with depression among the pregnant adolescents. This is at variance with findings among adults where unplanned pregnancy showed relationship with depression (43,44). This suggests that pregnancy in adolescence may be associated with emotional disturbance irrespective of the planned status or disposition of the adolescent to the pregnancy.

### Study strengths and Limitations

The strength of the study includes the focus on antenatal depression which has been less studied compared with postnatal depression. Furthermore, the inclusion of a comparison group and the use of structured instruments to assess the dependent and independent variables. However, there are few limitations, which include our inability to select the informal health facilities involved in this study randomly because effort made to get the sampling frame for the informal services was not successful. Also, screening instrument was used to assess depression as against structured diagnostic interview.

## Conclusion

This study concludes that the prevalence of depression is significantly higher among the pregnant adolescents compared with the non-pregnant adolescents. Living arrangement is the only socio-demographic factor that has a significant relationship with depression among pregnant adolescents while employment status in addition to living arrangement showed significant relationship among non-pregnant adolescents. On the other hand, history of mental illness, childhood sexual abuse and anxiety were the psychosocial factors with significant relationship with depression among pregnant adolescents and only anxiety among non-pregnant adolescents.

## Data Availability

Data set attached as supporting information in this submission

## References

1. Thapar A, Collishaw S, Pine DS, Thapar AK. Depression in adolescence. The Lancet. 2012;379(9820):1056–67.

2. Parker G, Brotchie H. Gender differences in depression. International Review of Psychiatry.2010 Oct 1;22(5):429–36.

3. Sajjadi H, Kamal SHM, Rafiey H, Vameghi M, Forouzan AS, Rezaei M. A Systematic Review of the Prevalence and Risk Factors of Depression among Iranian Adolescents. Glob J Health Sci.2013 May;5(3):16–27.

4. Field T, Diego M, Sanders C. Adolescent depression and risk factors. Adolescence. 2001;36(143):491–8.

5. WHO. Adolescent Pregnancy: Fact sheet. 2014; Available from: http://www.who.int.mediacentre/factsheets/fs364/en/

6. Mitsuhiro SS, Chalem E, Barros MM, Guinsburg R, Laranjeira R. Teenage pregnancy: use of drugs in the third trimester and prevalence of psychiatric disorders. Revista Brasileira de Psiquiatria. 2006;28:122–5.

7. Mitsuhiro SS, Chalem E, Moraes Barros MC, Guinsburg R, Laranjeira R. Brief report: Prevalence of psychiatric disorders in pregnant teenagers. Journal of Adolescence. 2009;32(3):747–52.

8. Marcus SM. Depression during pregnancy: rates, risks and consequences. Can J Clin Pharmacol. 2009;16(1):15–22.

9. Patel V, Rahman A, Jacob KS, Hughes M. Effect of maternal mental health on infant growth in low income countries: new evidence from South Asia. BMJ. 2004;328(7443):820–3.

10. Plant DT, Barker ED, Waters CS, Pawlby S, Pariante CM. Intergenerational transmission of maltreatment and psychopathology: the role of antenatal depression. Psychological Medicine. 2013;43(3):519–28.

11. Chalem E, Mitsuhiro SS, Manzolli P, Barros MCM, Guinsburg R, Sass N, et al. Underdetection of Psychiatric Disorders During Prenatal Care: A Survey of Adolescents in Sao Paulo, Brazil. Journal of Adolescent Health. 2012;50(1):93–6.

12. Adewuya AO, Ola BA, Aloba OO. Prevalence of major depressive disorders and a validation of the beck depression inventory among Nigerian adolescents. European Child and Adolescent Psychiatry. 2007;16(5):287–92.

13. Eisenberg D, Gollust SE, Golberstein E, Hefner JL. Prevalence and Correlates of Depression, Anxiety, and Suicidality Among University Students. American Journal of Orthopsychiatry.2007 Oct 1;77(4):534–42.

14. Eskin M, Ertekin K, Harlak H, Dereboy Ç. Prevalence of and Factors Related to Depression in High School Students. Turkish journal of psychiatry. 2008;19(4).

15. Sooky Z, Sharifi KH, Tagharrobi Z. The depression prevalence and its related factors in teenagers in Kashan-Iran 2006. European Psychiatry.2007 Mar 1;22:S246.

16. Unsal A, Ayranci U. Prevalence of Students With Symptoms of Depression Among High School Students in a District of Western Turkey: An Epidemiological Study. J School Health.2008 May;78(5):287–93.

17. Brown JD, Harris SK, Woods ER, Buman MP, Cox JE. Longitudinal study of depressive symptoms and social support in adolescent mothers. Maternal and Child Health Journal. 2012;16(4):894–901.

18. Coelho FM, Pinheiro RT, Silva RA, Quevedo Lde A, Souza LD, Castelli RD. Major depressive disorder during teenage pregnancy: socio-demographic, obstetric and psychosocial correlates. Revista brasileira de psiquiatria (Sao Paulo, Brazil: 1999) [Internet]. 2013;35. Available from: http://dx.doi.org/10.1016/j.rbp.2012.03.006

19. Nair M, Paul MK, John R. Prevalence of depression among adolescents. The Indian Journal of Pediatrics. 2004;71(6):523–4.

20. Logsdon MC, Cross R, Williams B, Simpson T. Prediction of Postpartum Social Support and Symptoms of Depression in Pregnant Adolescents: A Pilot Study. The Journal of School Nursing.2004 Feb 1;20(1):36–42.

21. Hoffmann F, Petermann F, Glaeske G, Bachmann CJ. Prevalence and Comorbidities of Adolescent Depression in Germany. Zeitschrift für Kinder-und Jugendpsychiatrie und Psychotherapie.2012 Oct 29;40(6):399–404.

22. Mojtabai R, Olfson M, Han B. National Trends in the Prevalence and Treatment of Depression in Adolescents and Young Adults. Pediatrics [Internet]. 2016 Dec 1 [cited 2019 Nov 15];138(6). Available from: https://pediatrics.aappublications.org/content/138/6/e20161878

23. Munhoz TN, Santos IS, Matijasevich A. Depression among Brazilian adolescents: A cross-sectional population-based study. Journal of Affective Disorders. 2015;175:281–6.

24. Rey JM, Sawyer MG, Clark JJ, Baghurst PA. Depression among Australian adolescents. Medical Journal of Australia. 2001;175(1):19–23.

25. Kim THM, Connolly JA, Tamim H. The effect of social support around pregnancy on postpartum depression among Canadian teen mothers and adult mothers in the maternity experiences survey. BMC Pregnancy and Childbirth. 2014;14(1):162–162.

26. Siegel RS, Brandon AR. Adolescents, Pregnancy, and Mental Health. Journal of Pediatric and Adolescent Gynecology. 2014;27(3):138–50.

27. Freitas GVS, Cais CFS, Stefanello S, Botega NJ. Psychosocial conditions and suicidal behavior in pregnant teenagers. European Child & Adolescent Psychiatry. 2008;17(6):336–336.

28. Biello KB, Sipsma HL, Kershaw T. Effect of Teenage Parenthood on Mental Health Trajectories: Does Sex Matter? American Journal of Epidemiology. 2010;172(3):279–87.

29. Troutman BR, Cutron CE. Nonpsychotic postpartum depression among adolescent mothers. J Abnorml Psychol [Internet]. 1990;99. Available from: https://doi.org/10.1037/0021-843X.99.1.69

30. Charan J, Biswas T. How to calculate sample size for different study designs in medical research? Indian journal of psychological medicine. 2013;35(2):121.

31. Steer RA, Scholl TO, Beck AT. Revised Beck Depression Inventory Scores of Inner-City Adolescents: Pre- and Postpartum. Psychol Rep.1990 Feb;66(1):315–20.

32. Kooiman CG, Ouwehand AW, Kuile MM. The Sexual and Physical Abuse Questionnaire (SPAQ) A screening instrument for adults to assess past and current experiences of abuse. Child abuse & neglect. 2002;26:939–53.

33. Folayan MO, Haire B, Harrison A, Odetoyingbo M, Fatusi O, Brown B. Ethical issues in adolescents’ sexual and reproductive health research in Nigeria. Developing world bioethics. 2015;15(3):191–8.

34. Wiemann CM, Rickert VI, Berenson AB, Volk RJ. Are pregnant adolescents stigmatized by pregnancy? J Adolesc Health.2005 Apr;36(4):352.e1-8.

35. Barnet B, Liu J, DeVoe M. Double jeopardy: Depressive symptoms and rapid subsequent pregnancy in adolescent mothers. Archives of Pediatrics & Adolescent Medicine. 2008;162(3):246–52.

36. Hallfors DD, Waller MW, Ford CA, Halpern CT, Brodish PH, Iritani B. Adolescent Depression and Suicide Risk Association with Sex and Drug Behavior. American Journal of Preventive Medicine 2004. 2004;27(3).

37. Villanueva LA, Pérez-Fajardo Mm, Fernando LI. [Sociodemographic factors associated with depression in pregnant adolescents]. Ginecol Obstet Mex.2000 Apr;68:143–8.

38. Monroe SM, Harkness KL. Is depression a chronic mental illness? Psychological Medicine.2012 May;42(5):899–902.

39. Lundberg P, Rukundo G, Ashaba S, Thorson A, Allebeck P, Östergren P-O, et al. Poor mental health and sexual risk behaviours in Uganda: A cross-sectional population-based study. BMC Public Health. 2011;11(1):125–125.

40. Noll JG, Shenk CE, Putnam KT. Childhood Sexual Abuse and Adolescent Pregnancy: A Meta-analytic Update. Journal of Pediatric Psychology. 2009;34(4):366–78.

41. Fergusson DM, Horwood LJ, Lynskey MT. Childhood Sexual Abuse and Psychiatric Disorder in Young Adulthood: II. Psychiatric Outcomes of Childhood Sexual Abuse. Journal of the American Academy of Child & Adolescent Psychiatry.1996 Oct 1;35(10):1365–74.

42. Weiss EL, Longhurst JG, Mazure CM. Childhood Sexual Abuse as a Risk Factor for Depression in Women: Psychosocial and Neurobiological Correlates. American Journal of Psychiatry.1999 Jun;156(6):816–28.

43. Faisal-Cury A, Menezes PR, Quayle J, Matijasevich A. Unplanned pregnancy and risk of maternal depression: secondary data analysis from a prospective pregnancy cohort. Psychology, Health & Medicine.2017 Jan 2;22(1):65–74.

44. Ege E, Timur S, Zincir H, Geçkil E, Sunar-Reeder B. Social support and symptoms of postpartum depression among new mothers in Eastern Turkey. Journal of Obstetrics and Gynaecology Research. 2008;34(4):585–93.

